# Outcomes of COVID-19 related hospitalisation among people with HIV in the ISARIC WHO Clinical Characterisation Protocol UK Protocol: prospective observational study

**DOI:** 10.1101/2020.08.07.20170449

**Authors:** Anna Maria Geretti, Alexander J. Stockdale, Sophie H. Kelly, Muge Cevik, Simon Collins, Laura Waters, Giovanni Villa, Annemarie Docherty, Ewen M Harrison, Lance Turtle, Peter JM Openshaw, J Kenneth Baillie, Caroline A. Sabin, Malcolm G Semple, Daniel Bradshaw, Alison Brown, Nicky Connor, Valerie Delpech, Saye Khoo, Tamyo Mbisa, Chloe Orkin, Ann Sullivan, ISARIC4C Investigators [Listed separately]

**Author notes:** **Correspondence**, For queries related to people with HIV, Prof Anna Maria Geretti, MD, PhD, FRCPath, Institute of Infection, University of Liverpool, 8 West Derby Street, Liverpool L69 7BE, For queries related to the ISARIC CCP-UK study, Prof Malcolm G Semple, PhD, FRCPE, FRCPCH, Institute of Infection, University of Liverpool, 8 West Derby Street, Liverpool L69 7BE. Joint first authors. Joint senior authors. CHASE study group (Daniel Bradshaw).

## Abstract

**Background.:** There is conflicting evidence about how HIV infection influences COVID-19. We compared the presentation characteristics and outcomes of people with and without HIV hospitalised with COVID-19 at 207 centres across the United Kingdom.

**Methods.:** We analysed data from people with laboratory confirmed or highly likely COVID-19 enrolled into the ISARIC CCP-UK study. The primary endpoint was day-28 mortality after presentation. We used Kaplan-Meier methods and Cox regression to describe the association with HIV status after adjustment for sex, ethnicity, age, indeterminate/probable hospital acquisition of COVID-19 (definite hospital acquisition excluded), presentation date, and presence/absence of ten comorbidities. We additionally adjusted for disease severity at presentation as defined by hypoxia/oxygen therapy.

**Findings.:** Among 47,539 patients, 115 (0·24%) had confirmed HIV-positive status and 103/115 (89·6%) had a record of antiretroviral therapy. At presentation, relative to the HIV-negative group, HIV-positive people were younger (median 55 versus 74 years; p<0·001), had a higher prevalence of obesity and moderate/severe liver disease, higher lymphocyte counts and C-reactive protein, and more systemic symptoms. The cumulative incidence of day-28 mortality was 25·2% in the HIV-positive group versus 32·1% in the HIV-negative group (p=0·12); however, stratification for age revealed a higher mortality among HIV-positive people aged below 60 years. The effect of HIV-positive status was confirmed in adjusted analyses (adjusted hazard ratio [HR] 1·49, 95% confidence interval [CI] 0·99-2·25; p=0·06). Following additional adjustment for disease severity at presentation, mortality was higher in HIV-positive people (adjusted HR 1·63; 95% CI 1·07-2·48; p=0·02). In the HIV-positive group, mortality was more common among those who were slightly older and among people with obesity and diabetes with complications.

**Interpretation.:** HIV-positive status may be associated with an increased risk of day-28 mortality following a COVID-19 related hospitalisation.

**Funding.:** NIHR, MRC, Wellcome Trust, Department for International Development, Bill and Melinda Gates Foundation.

**Study registration:** ISRCTN66726260

**RESEARCH IN CONTEXT:** *Evidence before this study:* We searched PubMed for articles in all languages containing the words “COVID*”, “coronavirus”, “SARS CoV-2” AND “HIV”. After screening on 23^rd^ July 2020, we found 51 articles reporting outcomes of COVID-19 in HIV-positive people. Of these, 2 were systematic reviews, 24 were single case reports or case series of under 10 participants, and 12 were larger case series or retrospective cohorts without matched controls. There were two cohort studies that matched HIV-positive people diagnosed with COVID-19 to the general population attending for HIV care in the same area, and three studies that matched HIV-positive people diagnosed with COVID-19 to HIV-negative controls. Some of the evidence from the United States and Europe to date suggests that people with HIV experience a similar disease course and outcomes of COVID-19 compared to the general population. However, many of the studies are limited by small sample size, lack of comparator group and lack of adjustment for potential confounding. In contrast, preliminary results from a cohort study of over 20,000 participants in South Africa indicate that HIV-positive status more than doubles the risk of COVID-19 related mortality. Currently, the evidence from the United Kingdom is limited to two case series comprising a total of 21 patients.

*Added value of this study:* This study analysed data collected from 207 sites across the United Kingdom as part of ISARIC CCP, the largest prospective cohort of patients hospitalised with COVID-19, to evaluate the association between HIV-positive status and day-28 mortality. The study has the benefit of a relatively large number of participants with HIV (n=115, almost all receiving antiretroviral therapy) and importantly, the ability to direct compare their presenting characteristics and outcomes to those of 47,424 HIV-negative controls within the same dataset. This includes the ability to assess the influence of gender, ethnicity and age, as well as the effect of key comorbidities including chronic cardiac, pulmonary, renal and haematological disease, diabetes, obesity, chronic neurological disorder, dementia, liver disease, and malignancy. Unlike some of the other evidence to date, but in line with the data from South Africa, this study indicates that HIV-positive status may increase the risk of mortality with COVID-19 compared to the general population, with an effect that was especially evident among people with HIV aged below 60 years and was independent of gender or ethnicity. Although we detected an association between mortality among people with HIV and occurrence of obesity and diabetes with complication, the effect of HIV-positive status persisted after adjusting for comorbidities.

*Implications of all the available evidence:* People with HIV may be at increased risk of severe outcomes from COVID-19 compared to the general population. Ongoing data collection is needed to confirm this association. Linkage of hospital outcome data to the HIV history will be paramount to establishing the determinants of the increased risk. COVID-19 related hospitalisation should pursue systematic recording of HIV status to ensure optimal management and gathering of evidence.

## Introduction

Factors associated with COVID-19 related mortality include older age and presence of chronic comorbidities, particularly obesity, chronic kidney disease (CKD), chronic obstructive pulmonary disease (COPD), serious cardiovascular disease, type II diabetes, and transplant-related immunosuppression.^1-5^ There is no conclusive evidence about the relationship with HIV infection. If untreated, HIV causes progressive immunosuppression; however, antiretroviral therapy (ART) restores immune function and life-expectancy.^6^ Immune restoration is not always complete despite effective ART, however, and a subset of people with HIV (PWH) remains at risk of persistent immune dysfunction,^7^ which might augment severity of COVID-19, or conversely, possibly reduce the immune responses that can complicate COVID-19.^8^ Although some antiretroviral drugs have been proposed to protect against COVID-19, the evidence remains uncertain.^9,10^ Importantly, HIV might increase the risk of adverse COVID-19 outcomes due to the common prevalence of co-factors such as CKD, COPD, and diabetes,^11^ alongside socioeconomic variables that may carry a negative influence.^12^

Several case series and observational cohort studies have described the outcomes of COVID-19 in PWH across Europe,^9,13-19^ Asia,^18,19^ and the United States.^8,18-22^ These studies have often been limited by small sample size, lack of direct comparative data from people without HIV, or inability to adjust for comorbidities. Some reports from Italy and New York indicated that HIV did not increase the risk of COVID-19 related hospitalisation or mortality,^4,14,21^ whereas two others suggested an increased risk of mortality among PWH hospitalised with COVID-19.^9,20^ Preliminary data from South Africa similarly suggest that HIV-positive status more than doubled the risk of COVID-19 related mortality.^23^

To characterise the presenting characteristics and outcomes of COVID-19 related hospitalisation in PWH relative to those without HIV in the United Kingdom (UK), we analysed data collected within the International Severe Acute Respiratory and emerging Infections Consortium (ISARIC) Clinical Characterisation Protocol (CCP), the largest prospective observational study of patients admitted to hospital with COVID-19 worldwide.^24^

## Methods

### Study design

ISARIC CCP-UK is an ongoing prospective cohort study in acute care hospitals in England, Scotland, and Wales.^24^ The protocol, case report form (CRF, version 9.2) and other study materials, and details of the Independent Data and Material Access Committee are available online.^24^ The study was activated in the UK on 17^th^ January 2020. Inclusion criteria were people aged ≥18 years who were admitted to one of the participating acute care trusts (207 at the time of data extraction) with either laboratory-confirmed or highly likely (based on clinical, laboratory and radiological findings) SARS CoV-2 infection. PCR-based virus detection in nasopharyngeal swabs was the only test available during the study and the decision to test was at the discretion of the attending clinical team, who also decided upon hospital admission, transfer into critical care and use of ventilation. For the present analyses, baseline was defined as the date of hospital admission or symptom onset (for those with symptom onset after hospitalisation, see below). Our analyses included individuals with a baseline date that was on or before 4^th^ June 2020 for whom ≥14 days had elapsed until the date of data extraction on 18^th^ June 2020. Individuals without information on the date of admission or with a baseline date after 4^th^ June 2020 were excluded. Where the date of symptom onset was missing, we assumed that symptoms began on the date of the SARS-COV-2 PCR test, or if this was not recorded, the date of admission. Information on positive HIV status, as reported to ISARIC CCP-UK, was confirmed through cross-checking with reported receipt of ART (n=103), receipt of *Pneumocystis jirovecii* prophylaxis in the absence of non-HIV indications (n=2), or directly with a site investigator (n=10). Individuals with missing HIV status and those with unconfirmed HIV-positive status were excluded from the analyses.

### Statistical analysis

Presenting characteristics were compared between HIV-positive and HIV-negative people and between PWH who died and those who survived to discharge using Wilcoxon rank sum tests (for continuous variables) and Pearson’s chi-squared or Fisher’s exact test (for categorical variables). For all individuals, follow-up ended on the date of death. Patients discharged to receive palliative care in the community were considered to have died three days following discharge. Follow-up was right censored at day 28 for those remaining alive as an inpatient, or for those who were discharged not for palliative care prior to day 28. A data check showed that the vast majority of those discharged prior to day 28 were still alive on this date. Follow-up on patients transferred to another facility was censored on the date of transfer. For patients who died, were transferred or discharged on the date of admission or who had no further follow-up recorded beyond the first day, we recorded 05 days of follow-up time. The primary analysis used a Kaplan-Meier approach to visually display the cumulative incidence of mortality over this period, overall and in strata defined by sex and age. Cox proportional hazards regression with the Efron method for ties was then used to describe the association of mortality with HIV status, before and after adjustment for the following potential confounders: sex, ethnicity, age (in quadratic form), indeterminate/probable hospital acquisition of COVID-19 (as defined above), and ten comorbidities at admission (a series of binary variables to indicate the presence or absence of each of chronic cardiac disease, chronic pulmonary disease, chronic renal disease, diabetes, obesity, chronic neurological disorder, dementia, liver disease [mild, moderate or severe], malignancy, and chronic haematological disease). We also included adjustment for the baseline date to account for changes in mortality over the period of interest. Where entries on comorbidity (presence or absence) were partially missing from the study CRF, we assumed that missing data indicated the absence of the specific comorbidity; however, participants with missing entries on all comorbidities were excluded from these adjusted analyses. Finally, we fitted a further model with additional adjustment for hypoxia at presentation, defined as oxygen saturation (SpO2) <94% on air or a record of receiving oxygen, as a marker of presenting disease severity, in order to assess whether any increased/decreased risk of mortality in PWH could be explained by a different stage of disease advancement at hospitalisation. A series of sensitivity analyses were performed for the main mortality outcome: i) we repeated the analyses after censoring follow-up on the day of discharge for those discharged before day 28; ii) we included those with definite hospital-acquired COVID-19 (baseline = date of symptom onset); iii) we used symptom onset date as the baseline date for all (rather than admission date where applicable); iv) we excluded PWH lacking a record of ART; v) we calculated propensity scores for HIV-positive status using a logistic regression model based on sex, ethnicity, age (in quadratic form), indeterminate/probable hospital acquisition of COVID-19, smoking status, baseline date, and ten comorbidities, and included the propensity score in a Cox regression model for death at 28 days; and vi) we considered a binary endpoint of 14-day mortality and performed logistic regression (with the same confounder adjustment as described above). In the HIV-positive group, we used a Cox proportional hazard model to investigate the associations of presenting characteristics with day-28 mortality. Analyses were conducted in Stata v161 (Statcorp, College Station, TX, USA).

## Results

### Participants

ISARIC CCP-UK recorded 53,992 people with COVID-19 between 17^th^ January 2020 and 18^th^ June 2020. After excluding non-eligible participants (Figure 1), the final analysis included 47,539 patients, of whom 115 (024%) had confirmed HIV-positive status. The characteristics of patients excluded from the analysis did not differ by sex, ethnicity or age; in particular, the characteristics of those excluded due to missing data on HIV status closely resembled those reported to be HIV-negative (Supplementary Table 1). Among PWH, one person was diagnosed with HIV during the admission and 103 (896%) had an ART record. The regional distribution of study participants with HIV compared to the total UK population of people accessing HIV care (2018 data) is shown in Supplementary Table 2.

**Figure 1.**
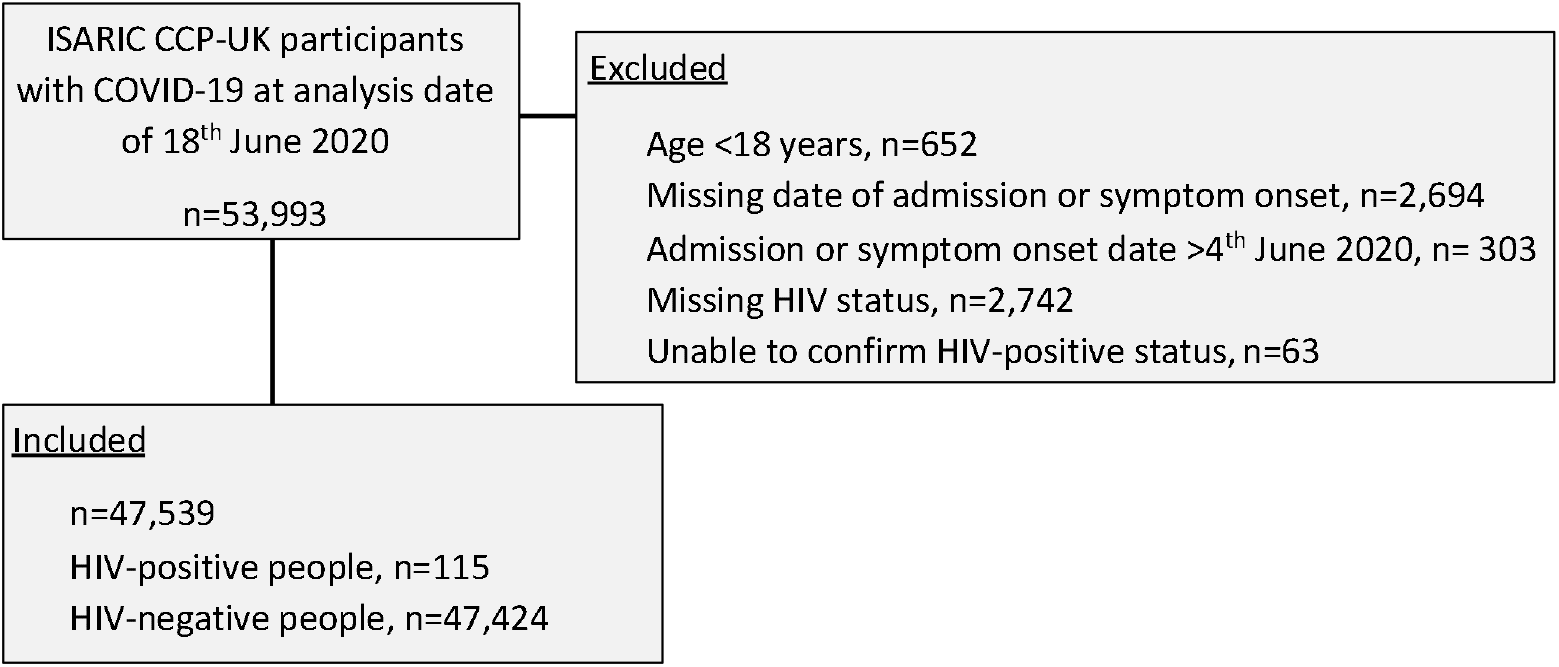
Flowchart of study participants.

### Characteristics at presentation

The presenting characteristics according to HIV status are summarized in Tables 1-3 and Figure 2. PWH were younger than HIV-negative people (medians of 55 versus 74 years, p<0001) (Table 1, Figure 2). There were fewer women in the HIV-positive group but significantly larger proportions of people of black ethnicity. A similar proportion had no recorded comorbidities, whereas occurrence of 2 comorbidities was more prevalent in the HIV-negative group. PWH had lower prevalence of chronic cardiac disease, chronic pulmonary disease, chronic neurological disorders, dementia, malignancy and rheumatological disease, and higher rates of obesity and moderate/severe liver disease. There were small differences in the prevalence of asthma, diabetes without complications, mild liver disease, and malnutrition, whereas proportions with CKD, diabetes with complications and chronic haematological disease were similar in the two groups.

**Table 1.** Summary of participant characteristics, stratified by HIV status

| Characteristic |  | HIV-positive<br>n=115 |  | HIV-negative<br>n=47,424 |  | P-value |
| --- | --- | --- | --- | --- | --- | --- |
| Age, median years (IQR) |  | 55 | (49, 61) | 74 | (60, 84) | <0.001 |
| Age group, n (%) | <40 | 7/113 | (6.2) | 2,574/46,882 | (5.5) | <0.001 |
|  | 40-49 | 26/113 | (23.0) | 3,242/46,882 | (6.9) |  |
|  | 50-59 | 47/113 | (41.6) | 5,940/46,882 | (12.7) |  |
|  | 60-69 | 23/113 | (20.4) | 7,266/46,882 | (15.5) |  |
|  | ≥70 | 10/113 | (8.9) | 27,860/46,882 | (59.4) |  |
| Female, n (%) |  | 39/114 | (34.2) | 20,280/47,258 | (42.9) | 0.06 |
| Ethnicity, n (%) | White | 44/106 | (41.5) | 35,501/42,163 | (84.2) | <0.001 |
|  | Black | 48/106 | (45.3) | 1,473/42,163 | (3.5) |  |
|  | Asian | 1/106 | (0.9) | 2,247/42,163 | (5.3) |  |
|  | Other | 13/106 | (12.3) | 2,942/42,163 | (7.0) |  |
| Smoking, n (%) | Never | 62/89 | (69.7) | 17,380/30,348 | (57.3) | 0.002 |
|  | Former | 16/89 | (18.0) | 10,632/30,348 | (35.0) |  |
|  | Current | 11/89 | (12.4) | 2,336/30,348 | (7.7) |  |
| Comorbidities, median number (IQR) |  | 1 | (0, 2) | 2 | (1, 3) | <0.001 |
| Comorbidities, n (%) | None | 30/115 | (26.1) | 9,675/46,696 | (20.7) | 0.002 |
|  | 1 | 44/115 | (38.3) | 13,532/46,696 | (29.0) |  |
|  | 2 | 29/115 | (25.2) | 11,518/46,696 | (25.2) |  |
|  | ≥3 | 12/115 | (10.4) | 11,971/46,696 | (25.6) |  |
| Type of comorbidities, n (%) | Chronic cardiac disease | 20/111 | (18.0) | 14,603/45,008 | (32.5) | 0.001 |
|  | Chronic pulmonary disease<sup>a</sup> | 12/114 | (10.5) | 8,048/44,872 | (17.9) | 0.04 |
|  | Asthma | 11/110 | (10.0) | 6,228/44,712 | (13.9) | 0.23 |
|  | Chronic renal disease | 19/110 | (17.3) | 7,861/44,683 | (17.6) | 0.93 |
|  | Diabetes, no complications | 16/110 | (14.6) | 7,776/43,818 | (17.8) | 0.38 |
|  | Diabetes, with complications | 8/110 | (7.3) | 3,300/43,543 | (7.6) | 0.90 |
|  | Obesity | 19/105 | (18.1) | 4,588/40,419 | (11.4) | 0.03 |
|  | Chronic neurological disorder | 7/110 | (6.4) | 5,579/44,432 | (12.6) | 0.05 |
|  | Dementia | 3/111 | (2.7) | 7,458/44,508 | (16.8) | <0.001 |
|  | Mild liver disease | 3/111 | (2.7) | 631/44,183 | (1.4) | 0.21 |
|  | Moderate/severe liver disease | 6/111 | (5.4) | 858/43,378 | (1.9) | 0.008 |
|  | Malignancy | 3/111 | (2.7) | 4,586/44,314 | (10.4) | 0.004 |
|  | Chronic haematological disease | 4/111 | (3.6) | 1,923/44,266 | (4.3) | 1.0 |
|  | Rheumatological disease | 6/111 | (5.4) | 4,867/44,138 | (11.0) | 0.06 |
|  | Malnutrition | 5/105 | (4.8) | 1,129/41,811 | (2.7) | 0.21 |

<sup>a</sup>Excludes asthma. Abbreviations: IQR, Interquartile range.

**Table 2.** Presenting symptoms and observations, stratified by HIV status

| Symptoms and Observations |  | HIV-positive<br>n=115 | HIV-negative<br>n=47,424 | P-value |
| --- | --- | --- | --- | --- |
| Presenting symptoms, n (%) | Fever | 94/114 (82.5) | 30,623/47,039 (65.1) | <0.001 |
|  | Myalgia | 26/96 (27.1) | 6,349/34,806 (18.2) | 0.03 |
|  | Headache | 18/89 (20.2) | 3,660/34,762 (10.5) | 0.003 |
|  | Cough | 88/114 (77.2) | 30,997/47,031 (65.9) | 0.01 |
|  | Dyspnoea | 84/114 (73.7) | 32,108/46,997 (68.3) | 0.22 |
|  | Chest pain | 24/101 (23.8) | 5,221/38,269 (13.6) | 0.003 |
|  | Sore throat | 13/92 (14.1) | 2,803/34,265 (8.2) | 0.04 |
|  | Wheeze | 5/94 (5.3) | 3,289/36,209 (9.1) | 0.28 |
|  | Rhinorrhoea | 3/89 (3.4) | 831/33,533 (2.5) | 0.49 |
|  | Diarrhoea | 25/100 (25.0) | 7,271/39,305 (18.5) | 0.10 |
|  | Nausea or vomiting | 23/97 (23.7) | 7,641/39,527 (19.3) | 0.28 |
|  | Abdominal pain | 13/96 (13.5) | 4,026/38,105 (10.6) | 0.34 |
|  | Fatigue | 40/90 (44.4) | 16,098/37,169 (43.3) | 0.83 |
|  | Asymptomatic | 0/115 (0) | 887/47,424 (1.9) | 0.28 |
| Symptom group<sup>a</sup>, n (%) | Systemic | 103/114 (90.4) | 32,239/47,073 (68.5) | <0.001 |
|  | Respiratory | 100/114 (87.7) | 38,697/47,112 (82.1) | 0.12 |
|  | Gastrointestinal | 42/103 (40.8) | 13,431/41,275 (32.5) | 0.08 |
| Symptom duration, median days (IQR) |  | 5 (1, 9) | 3 (0, 7) | 0.001 |
| Symptom duration<sup>b</sup>, n (%) | <3 days | 108/115 (93.9) | 41,247/47,055 (87.7) | 0.13 |
|  | 3-7 days | 1/115 (0.9) | 1,484/47,055 (3.2) |  |
|  | 8-14 days | 4/115 (3.5) | 1,602/47,055 (3.4) |  |
|  | >14 days | 2/115 (1.7) | 2,722/47,055 (5.8) |  |
| Presenting signs | Temperature, median °C (IQR) | 37.8 (37.0, 38.6) | 37.3 (36.6, 38.1) | 0.002 |
|  | Fever ≥37.8 °C, n (%) | 59/111 (53.2) | 16,432/45,414 (36.2) | <0.001 |
|  | HR, median beats/min (IQR) | 97 (82, 110) | 90 (78, 105) | 0.002 |
|  | Tachycardia<sup>c</sup>, n (%) | 51/112 (45.5) | 15,064/45,387 (33.2) | 0.006 |
|  | RR, median breaths/min (IQR) | 20 (18, 27) | 21 (18, 26) | 0.71 |
|  | Tachypnoea<sup>d</sup>, n (%) | 50/107 (46.7) | 23,284/45,165 (51.6) | 0.32 |
|  | Hypoxia<sup>e</sup>/on oxygen, n (%) | 50/108 (46.3) | 23,948/45,199 (53.0) | 0.16 |
|  | Infiltrates visible on CXR, n (%) | 46/70 (65.7) | 19,047/30,536 (62.4) | 0.57 |
|  | Systolic BP, median mmHg (IQR) | 130 (117, 143) | 130 (114, 147) | 0.90 |
|  | Diastolic BP, median mmHg (IQR) | 80 (68, 88) | 74 (65, 84) | 0.009 |

<sup>a</sup>Systemic symptoms: ≥1 of fever, myalgia or headache; Respiratory symptoms: ≥1 of cough, dyspnoea, chest pain, sore throat, wheeze; Gastrointestinal symptoms: ≥1 of: Diarrhoea, nausea, vomiting or abdominal pain. <sup>b</sup>Based on the onset of symptoms relative to the date of admission, COVID-19 acquisition was classed as community (<3 days), indeterminate (3-7 days), probable hospital (8-14 days), and definite hospital (>14 days). <sup>c</sup>Defined as HR >100 beats/min. <sup>d</sup>Defined as RR >20 breaths/min. <sup>e</sup>Defined as SpO<sub>2</sub> <94% on air; proportions with SpO<sub>2</sub> <94% or on oxygen therapy at presentation were 30/108 (27.8%) and 32/105 (30.5%) respectively in the HIV-positive group, and 14,463/45,123 (32.1%) and 14,203/44,093 (32.2%) in the HIV-negative group. Abbreviations: IQR, Interquartile range; HR, Heart rate; RR, Respiratory rate; CXR, Chest X-ray; BP, Blood pressure.

**Table 3.** Presenting laboratory parameters, stratified by HIV status

| Laboratory parameter | HIV positive<br>n=115 |  | HIV-negative<br>n=47,424 |  | P-value |
| --- | --- | --- | --- | --- | --- |
| Haemoglobin, median g/L (IQR) | 129 | (116, 144) | 129 | (113, 143) | 0.77 |
| Anaemia<sup>a</sup>, n (%) | 37/101 | (36.6) | 15,549/40,450 | (38.8) | 0.65 |
| WBC, median count x10<sup>9</sup>/L (IQR) | 6.6 | (4.9, 9.0) | 7.4 | (5.4, 10.4) | 0.01 |
| Lymphocytes, median count x10<sup>9</sup>/L (IQR) | 1.0 | (0.8, 1.5) | 0.9 | (0.6, 1.3) | <0.001 |
| Lymphopenia<sup>b</sup>, n (%) | 47/102 | (46.1) | 22,991/39,719 | (57.9) | 0.02 |
| Platelets, median count x10<sup>6</sup>/L (IQR) | 200 | (150, 263) | 217 | (164, 285) | 0.04 |
| Thrombocytopenia<sup>c</sup>, n (%) | 24/99 | (24.2) | 7,431/39,698 | (18.7) | 0.16 |
| Prothrombin time, median sec (IQR) | 13.6 | (11.0, 15.0) | 13.2 | (11.8, 15.0) | 0.71 |
| Creatinine, median µmol/L (IQR) | 89 | (72, 134) | 86 | (67, 121) | 0.21 |
| eGFR<sup>d</sup>, median ml/min/1.73m<sup>2</sup> (IQR) | 76 | (52, 101) | 73 | (48, 97) | 0.30 |
| eGFR ml/min/1.73m<sup>2</sup>,<br>n (%) |  |  |  |  | 0.15 |
| ≥60 | 67/95 | (70.5) | 24,758/38,782 | (63.8) |  |
| 30-59 | 18/95 | (19.0) | 9,774/38,782 | (25.2) |  |
| <30 | 10/95 | (10.5) | 4,250/38,782 | (11.0) |  |
| 15-29 | 4/95 | (4.2) | 2,844/38,782 | (7.3) |  |
| <15 | 6/95 | (6.3) | 1,406/38,782 | (3.6) |  |
| ALT, median U/L (IQR) | 27 | (19, 44) | 26 | (17, 43) | 0.28 |
| ALT >40 U/L, n (%) | 26/85 | (30.6) | 8,452/30,443 | (27.8) | 0.56 |
| Glucose, median mmol/L (IQR) | 6.7 | (5.8, 9.8) | 6.8 | (5.8, 8.9) | 0.63 |
| Hyperglycaemia<sup>e</sup>, n (%) | 10/52 | (19.2) | 2,900/19,522 | (14.9) | 0.38 |
| C-reactive protein, median mg/L (IQR) | 110 | (51, 200) | 83 | (36, 157) | 0.02 |

<sup>a</sup>Defined as haemoglobin <130 g/L in males and <115 g/L in females. <sup>b</sup>Defined as lymphocyte count <1.0 x10<sup>9</sup>/L. <sup>c</sup>Defined as platelet count <150 x10<sup>6</sup>/L. <sup>d</sup>Based on the Modification of Diet in Renal Disease (MDRD) formula where eGFR (mL/min/1.73 m<sup>2</sup>) = 175 × (Scr/88.4)<sup>-1.154</sup> × (Age)<sup>-0.203</sup> × (0.742 if female) × (1.212 if Black ethnicity). <sup>e</sup>Defined as glucose >11 mmol/L. Abbreviations: IQR, Interquartile range; Abbreviations: IQR, Interquartile range; WBC, White blood cells; eGFR, estimated glomerular filtration rate; ALT, alanine transaminase.

**Figure 2.**
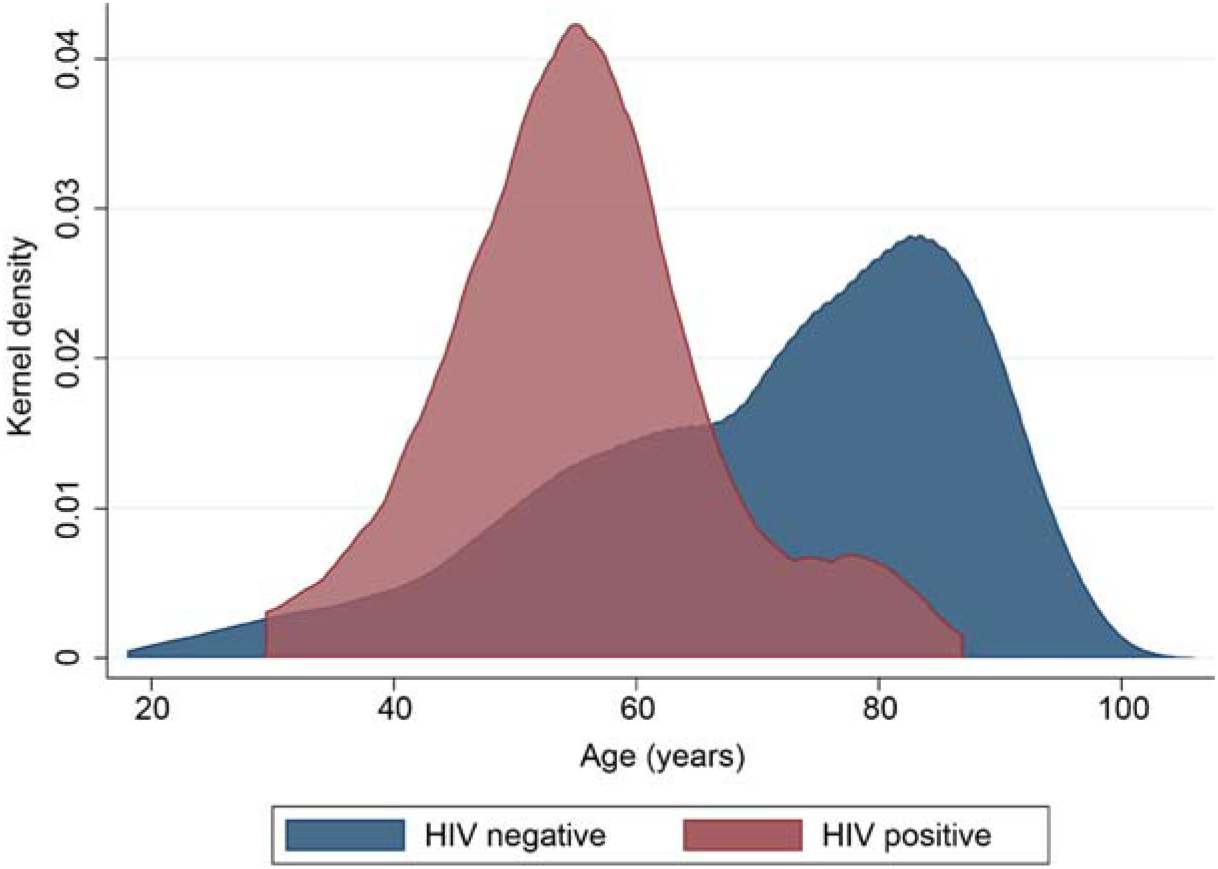
Kernel density plot of age distribution of study participants stratified by HIV status.

The duration of symptoms was longer in the HIV-positive group (medians of 5 vs. 3 days, p=0001) (Table 2). PWH were more likely to present with systemic symptoms and signs, including fever, headache, myalgia and tachycardia, and to have cough, sore throat and chest pain. To a lesser extent, they also had more common occurrence of gastrointestinal symptoms. Respiratory rate, occurrence of tachypnoea and hypoxia, and radiological evidence of chest infiltrates did not show significant differences between the two groups. PWH presented with lower total white blood cell and platelet count, but higher lymphocyte count and C-reactive protein (CRP) (Table 3). Other laboratory parameters showed no significant differences.

### COVID-19 outcomes

During admission, whereas there was no significant difference in the proportion of participants who received oxygen between the two groups, significantly higher proportions of PWH were admitted to critical care and received non-invasive and invasive ventilation (Figure 3). However, after adjustment for sex, ethnicity, age, baseline date, indeterminate/probable hospital acquisition of COVID-19, and ten comorbidities, the odds of admission to critical care were similar between the two groups (odds ratio [OR] 1·13; 95% confidence interval [CI] 0-72-1-75; p=059) (Supplementary Table 3).

**Figure 3.**
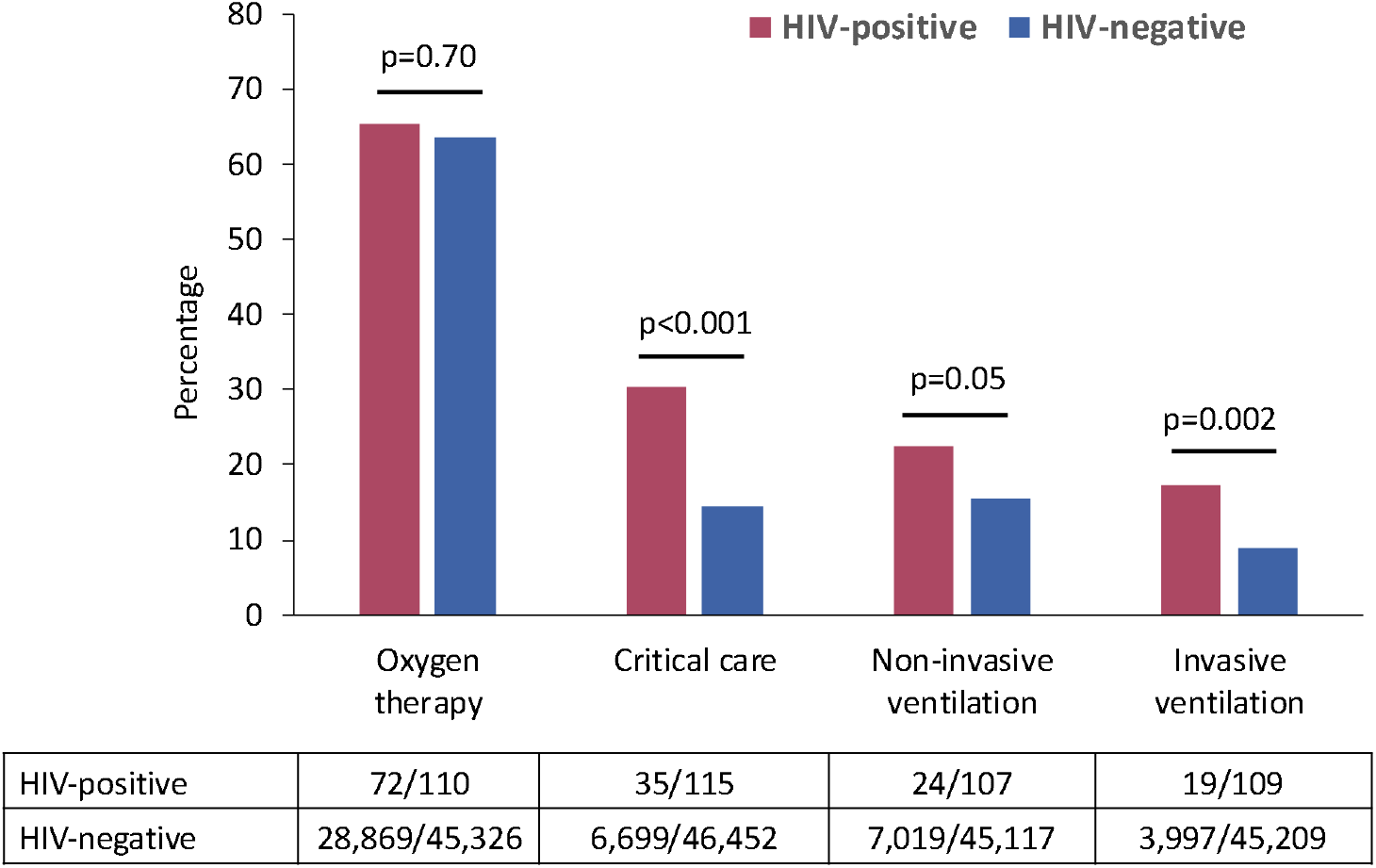
Interventions during hospitalisation by HIV status.

Overall, by day 28, 13,981 (29·4%) participants were known to have died, 23,642 (49·7%) had been discharged alive, 3,715 (7·8%) remained in hospital, and 1,801 (3·8%) had transferred to other facilities; in the remaining 4,400 (9·2%) the outcome was unknown (Supplementary Table 4). In the HIV-positive group, 26 (22·6%) were known to have died compared to 13,955 (29·1%) of the HIV-negative group; the cumulative incidence of day-28 mortality was 25·2% vs. 32·1%, respectively (p=0·12, log-rank test, Figure 4 A). Whilst findings were similar (in unadjusted analyses) regardless of sex (Figure 4 B, C), stratification for age revealed higher mortality among PWH in the two younger age groups (<50 years and 50-59 years) but not in those aged 60-79 years (Figure 3 D-F).

**Table 4.** Cox proportional hazards model of the association between HIV status and day-mortality

| HIV-positive versus HIV-negative | Hazard ratio | 95% CI | P-value |
| --- | --- | --- | --- |
| Unadjusted | 0.74 | 0.50-1.09 | 0.12 |
| Adjusted for sex | 0.71 | 0.48-1.05 | 0.08 |
| Adjusted for ethnicity | 0.77 | 0.52-1.15 | 0.21 |
| Adjusted for age | 1.39 | 0.94-2.09 | 0.10 |
| Adjusted for age and sex | 1.39 | 0.93-2.08 | 0.11 |
| Adjusted for sex, ethnicity, age, baseline date, and indeterminate/probable hospital acquisition of COVID-19 | 1.49 | 0.99-2.25 | 0.06 |
| Adjusted for sex, ethnicity, age, baseline date, indeterminate/probable hospital acquisition of COVID-19, and 10 comorbidities<sup>a</sup> | 1.49 | 0.99-2.26 | 0.06 |
| Adjusted for sex, ethnicity, age, baseline date, indeterminate/probable hospital acquisition of COVID-19, 10 comorbidities<sup>a</sup>, and hypoxia at presentation<sup>b</sup> | 1.62 | 1.06-2.46 | 0.02 |
| Adjusted for sex, ethnicity, age, baseline date, indeterminate/probable hospital acquisition of COVID-19, 10 comorbidities<sup>a</sup> and hypoxia/ receiving oxygen at presentation<sup>b</sup> | 1.63 | 1.07- 2.48 | 0.02 |

<sup>a</sup>The model adjusted for the following comorbidities: chronic cardiac disease, chronic pulmonary disease, chronic renal disease, diabetes, obesity, chronic neurological disorder, dementia, liver disease, malignancy, and chronic haematological disease. <sup>b</sup>Hypoxia was defined as SpO<sub>2</sub> <94% on air; a record of hypoxia or receiving oxygen at presentation were used as an indicator of disease severity.

**Figure 4.**
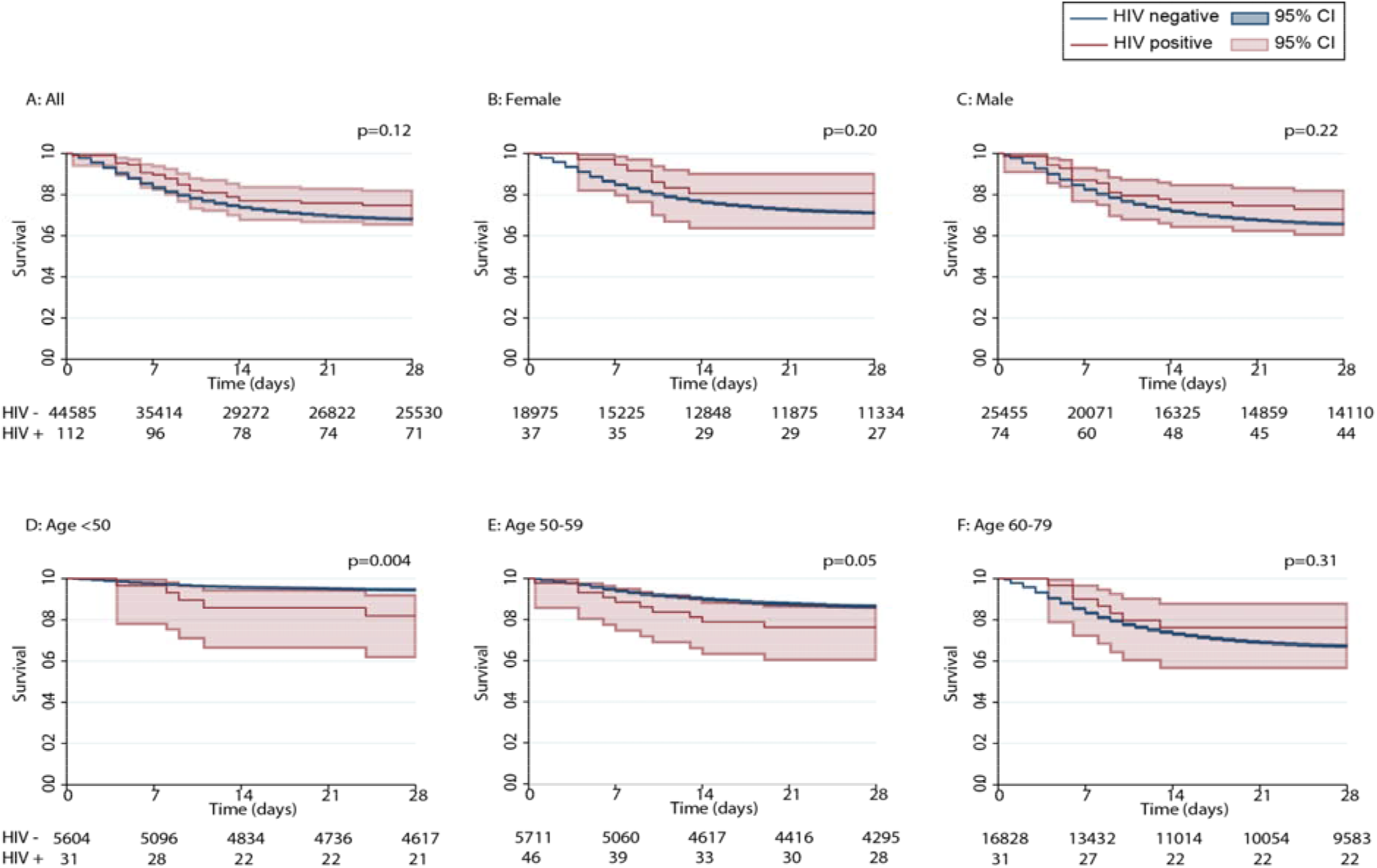
Kaplan Meier survival graphs, stratified by HIV status, sex and age group. P values represent log-rank tests. Plots D, E and F include only individuals from age groups <50 years, 50-59 years and 60-79 years.

In unadjusted models (Table 4), the cumulative hazard of day-28 mortality was 26% lower in HIV-positive vs. HIV-negative people (HR 074, 95% CI 0·50-1·08; p=0·12). The hazard was similar after adjustment for either sex or ethnicity; in contrast, and as expected based on the stratified Kaplan-Meier analyses, adjustment for age resulted in a change in the direction of the association (adjusted HR 1·39, 95% CI 0·94-2·09; p=0·10) (Table 4). After further adjustment for sex, ethnicity, age, baseline date, indeterminate/probable hospital acquisition of COVID-19 and ten co-morbidities, the risk of mortality was 49% higher in PWH (adjusted HR 1·49; 95% CI 0·99-2·26; p=0·06). Following additional adjustment for disease severity at presentation (based on a record of hypoxia or oxygen therapy), the risk of mortality was 63% higher in PWH (adjusted HR 1·63; 95% CI 1·07-2·48; p=0·02) (Table 4). After day 28, there were no deaths recorded in the HIV-positive group whereas 586 deaths occurred in the HIV-negative group.

Sensitivity analyses showed consistent results (Supplementary Table 5). In particular, censoring follow-up on the day of discharge for those discharged before day 28, including patients with definite hospital acquired COVID-19, using symptom onset as the start of follow-up, or excluding PWH lacking an ART record did not significantly alter the model. A separate logistic regression model with a binary variable of day-14 mortality showed increased odds of mortality in the HIV-positive group (adjusted OR 177; 95% CI 1·06-2·95; p=0·03).

In the HIV-positive group, relative to patients who survived by day 28, patients who died were slightly older and had a higher prevalence of obesity and diabetes with complications (Table 5 and Supplementary Tables 6 and 7). An ART record was more often missing among those who died. No indications of other major differences were observed.

**Table 5.** Characteristics of patients with HIV, stratified by outcome at day 28, selected variables^a^

| Characteristic | Died n=26 | Alive n=89 | P-value |
| --- | --- | --- | --- |
| Age, median years (IQR) | 58 (52, 72) | 54.5 (49, 60) | 0.07 |
| Age group, n (%) |  |  | 0.06 |
| <40 | 1/24 (4.2) | 6/89 (6.7) |  |
| 40-49 | 4/24 (16.7) | 22/89 (24.7) |  |
| 50-59 | 10/24 (41.7) | 37/89 (41.6) |  |
| 60-69 | 3/24 (12.5) | 20/89 (22.5) |  |
| ≥70 | 6/24 (25.0) | 4/89 (4.5) |  |
| ART recorded, n (%) | 22/26 (84.6) | 83/89 (93.3) | 0.17 |
| Type of comorbidities, n (%) |  |  |  |
| Chronic pulmonary disease<sup>b</sup> | 0/26 (0) | 12/88 (13.6) | 0.05 |
| Diabetes, with complications | 5/26 (19.2) | 3/84 (3.6) | 0.02 |
| Obesity | 8/24 (33.3) | 11/81 (13.6) | 0.03 |
| Presenting symptoms, n (%) |  |  |  |
| Cough | 22/25 (88.0) | 66/89 (74.2) | 0.15 |
| Diarrhoea | 8/22 (36.4) | 17/78 (21.8) | 0.16 |
| Symptom group, n (%) |  |  |  |
| Respiratory<sup>c</sup> | 24/25 (96.0) | 76/89 (85.4) | 0.15 |
| Presenting signs |  |  |  |
| HR, median beats/min (IQR) | 106 (96, 120) | 93 (80, 108) | <0.001 |
| Tachycardia<sup>d</sup>, n (%) | 16/25 (64.0) | 35/87 (40.2) | 0.04 |
| RR, median breaths/min (IQR) | 26 (19, 30) | 20 (18, 24) | 0.02 |
| Tachypnoea<sup>e</sup>, n (%) | 15/23 (65.2) | 35/84 (41.7) | 0.05 |
| Hypoxia<sup>f</sup>/on oxygen, n (%) | 18/24 (75.0) | 32/84 (38.1) | 0.001 |
| Diastolic BP, median mmHg (IQR) | 70 (62, 81) | 81 (69, 89) | 0.04 |
| WBC, median count x10<sup>9</sup>/L (IQR) | 7.8 (5.5, 10.9) | 5.7 (4.7, 8.7) | 0.02 |
| Laboratory parameters |  |  |  |
| eGFR<sup>g</sup>, median ml/min/1.73m<sup>2</sup> (IQR) | 72 (51, 87) | 78 (58, 102) | 0.12 |
| Glucose, median mmol/L (IQR) | 10.3 (6.4, 12.3) | 6.4 (5.8, 8.3) | 0.06 |
| Hyperglycaemia<sup>h</sup>, n (%) | 5/13 (38.5) | 5/39 (12.8) | 0.10 |
| C-reactive protein, median mg/L (IQR) | 203 (103, 306) | 92 (42, 149) | <0.001 |
| Interventions, n (%) |  |  |  |
| Oxygen therapy during admission | 21/26 (80.8) | 51/84 (60.7) | 0.06 |
| Critical care admission | 17/26 (65.4) | 18/89 (20.2) | <0.001 |
| Non-invasive ventilation | 9/24 (37.5) | 15/83 (18.1) | <0.001 |
| Invasive ventilation | 13/26 (50.0) | 6/88 (6.8) | <0.001 |

<sup>a</sup>A full list of demographic and clinical characteristics is shown in Supplementary Table 6. <sup>b</sup>Excludes asthma. <sup>c</sup>Respiratory symptoms: ≥1 of cough, dyspnoea, chest pain, sore throat, wheeze. <sup>d</sup>Defined as HR >100 beats/minute. <sup>e</sup>Defined as RR >20 breaths/min. <sup>f</sup>Defined as SpO<sub>2</sub> <94% on air. <sup>g</sup>Based on the Modification of Diet in Renal Disease (MDRD) formula where eGFR (mL/min/1.73 m<sup>2</sup>) = 175 × (Scr/88.4)<sup>-1.154</sup> × (Age)<sup>-0.203</sup> × (0.742 if female) × (1.212 if Black ethnicity). <sup>h</sup>Defined as glucose >11 mmol/L. Abbreviations: IQR, Interquartile range; ART, Antiretroviral therapy; HR, Heart rate; RR, Respiratory rate; BP, Blood pressure; WBC, White blood cells; eGFR, estimated glomerular filtration rate.

## Discussion

### Principal findings

In this study of 115 HIV-positive and 47,979 HIV-negative people, we found evidence suggesting a 63% increased risk of day-28 mortality among PWH hospitalised with COVID-19 compared to HIV-negative individuals in the same dataset, after adjustment for sex, ethnicity, age, baseline date, ten key comorbidities, and disease severity at presentation (as indicated by a record of hypoxia or receiving oxygen therapy). The latter adjustment took into consideration that doctors may be more likely to admit HIV-positive people with COVID-19 despite less severe symptoms. A striking difference in mortality was seen in the younger age groups (<50 years and 50-59 years), although the number of older PWH was small.

The role of age, sex and ethnicity on COVID-19 outcomes is the focus of much research.^1,2,25^ PWH in our study were significantly younger than the HIV-negative group and adjusting for age changed the direction of the association between HIV status and day-28 mortality, suggesting that age was a significant confounder in our analyses. Men were more prevalent in the HIV-positive group, which is consistent with the epidemiology of HIV infection in the UK, where men represent just over two thirds of the whole population with HIV.^26^ People ofblack ethnicity made up nearly 42% of the HIV-positive group, whereas men and women of black African ethnicity account for ~26% of the total number of PWH in the UK.^26^ Nonetheless, adjustment for sex or ethnicity alone did not impact our relative hazard estimates.

Whilst there is a recognised interplay between HIV and comorbidities, omitting the adjustment for comorbidities did not modify the association. PWH had fewer comorbidities, notably lower prevalence of chronic pulmonary disease and malignancies, and this is likely to be partly a function of their younger age. HIV-positive people who died were older and were more likely to suffer from obesity and diabetes with complications than those who survived to discharge. Similar trends have been seen in the general population.^1,25^ While these observations highlight the importance of obesity and diabetes as cofactors, adjustment for comorbidities in our model did not modify the association, suggesting that the apparent increased risk of COVID-19 related mortality in PWH was not merely due to the presence of promoting comorbidities.

### Comparison with other studies

Evidence from published studies is not entirely consistent about the interplay between HIV and COVID-19.^8,9,13-23^ A case-control study from New York compared 88 PWH, all of whom were receiving ART, and 405 HIV-negative controls matched by age, gender, ethnicity, and calendar week of infection.^21^ The study found no difference in the outcomes of COVID-19 related hospitalisation after adjusting for demographics, COPD, smoking, and baseline ferritin and white blood cell count. Apart from the differences in the study design and geography, there are fundamental differences between our study population and the New York cohort described by Sigel et al.^21^ Most importantly, participants in the latter study were older, with a median age of 61 years (IQR 54-67 years), whereas we found that the excess mortality occurred in HIV-positive people aged below 60 years. Whereas malignancies were recorded less commonly (3% vs. 10%), prevalence of obesity was nearly double in our cohort (18% vs. 11%). At the other spectrum, preliminary data from the Western Cape Department of Health in South Africa indicate that HIV-positive status was associated with increased hazard of mortality [adjusted HR 275].^23^ Although the South African model did not account for history of tuberculosis, obesity and socioeconomic status, it is of significance that HIV suppression on ART made no difference to the risk.

### Strengths and limitations of this study

A key strength of our study is the ability to perform a direct comparison of people with and without HIV in the same dataset. Our analysis does not address risk factors for a COVID-19 diagnosis or a COVID-19 related hospitalisation among PWH, and cannot add to the current debate about the role of certain antiretroviral agents in modulating such risks.^9,10^ In addition, due to the format of data collection in ISARIC CCP-UK, our analysis cannot provide evidence of the role of HIV-related parameters on outcomes of COVID-19 related hospitalisation, as we did not have details of the ART history, current and nadir CD4 cell count, plasma HIV-1 RNA load, and history of previous HIV-related disease. Only a subset of CRFs from participants with HIV included a record of receiving ART and the records were frequently incomplete.

Numerically, HIV-positive people who died were more likely not to have a record of being on ART than those who remained alive at day 28. However, it cannot be stated with certainty that those lacking an ART record were untreated nor therefore that lack of ART played a role in the adverse outcomes. Our experience of working with large HIV datasets is that we often find that people with missing data have worse mortality outcomes, simply because mortality prevents collection of a detailed treatment history. In the UK, 93% of the 103,000 people estimated to have HIV infection have been diagnosed and of these the vast majority (97%) receives ART and maintains excellent suppression of the infection.^26^ Among those with a HIV diagnosis, only a small subset of ~3% is either not engaged with care or experiences problems with virological control despite ART.^26^ This suggests that the likelihood of PWH in our study being off ART despite an absent record was overall low.

It is currently unclear how HIV infection and associated immune dysfunction modulates infection with SARS CoV-2. Whilst immunosuppression was associated with poor COVID-19 outcomes in a recent meta-analysis,^5^ effective ART leads to immune reconstitution with improved or normalised CD4 cell counts.^6^ We found no evidence of increased lymphopenia among PWH in our study. Furthermore, compared to HIV-negative people, PWH were more likely to experience systemic symptoms with fever and also showed higher CRP levels. These observations are likely to be reflective of the younger age of PWH in our study,^27^ and at the same time indicate preserved inflammatory responses in this group. This suggests that the likelihood of PWH in our study being severely immunosuppressed was overall low.

### Conclusions and policy Implications

After careful considerations and multiple adjustments for demographics, comorbidities and disease severity on admission, our initial analyses of the outcomes of patients hospitalised with COVID-19 in the UK show a signal towards an increased risk of day-28 mortality due to HIV-positive status. The data for this study were collected during the peak of the UK COVID-19 epidemic and the analysis contains a significant proportion of missing data, including a high number of patients with missing HIV status, who were excluded from the analysis. As the pandemic continues to spread in areas of increased HIV prevalence, our observations highlight the importance of recording the HIV status of people hospitalised with COVID-19 to ensure appropriate management during hospitalisation and gather further data to improve our understanding of the reciprocal interactions between SARS-CoV-2 and HIV.

Despite effective ART and normalised CD4 cell counts, a subset of PWH continue to experience immune activation, inflammation and a pro-coagulatory state,^7^ which may be postulated to modulate the risk of COVID-19 related morbidity and mortality.^28,29^ One determinant of such persistent immune dysfunction is the degree of immunosuppression experienced prior to the start of ART, defined by a low nadir CD4 cell count and inverted CD4:CD8 ratio. In the UK, 43% of people newly diagnosed with HIV in 2018 had a CD4 count <350 cells/mm^3^, a threshold indicative of significant immunosuppression.^26^ Furthermore, current guidelines about starting ART immediately at the time to diagnosis were implemented relatively recently, whereas in the past ART initiation was deferred until the CD4 count had declined below thresholds of initially 200, then 350 and subsequently 500 cells cells/mm^3^.^6,30^ Thus, many PWH in the UK and worldwide will have experienced years of uncontrolled HIV replication prior to commencing treatment, and may have experienced earlier regimens of suboptimal efficacy, with lasting effects on immune function. Planned linkage of the hospital dataset with the HIV clinic records will be required to clarify the role of ART history, current and nadir CD4 cell count, plasma HIV-1 RNA load and previous history of HIV-related disease on the outcomes observed in this study. Meanwhile, emphasis for PWH should be placed on early HIV diagnosis, prompt ART initiation, and optimised screening for and control of comorbidities including obesity and diabetes.

## Data Availability

The CO-CIN data were collated by ISARIC4C Investigators. ISARIC4C welcomes applications for data and material access through our Independent Data and Material Access Committee (https://isaric4c.net).

## ETHICAL CONSIDERATIONS

Ethical approval was given by the South Central - Oxford C Research Ethics Committee in England (Ref 13/SC/0149), the Scotland A Research Ethics Committee (Ref 20/SS/0028), and the WHO Ethics Review Committee (RPC571 and RPC572, 25 April 2013

## ACKNOWLEDGEMENTS

This work uses data provided by patients and collected by the NHS as part of their care and support #DataSavesLives. We are extremely grateful to the 2,648 frontline NHS clinical and research staff and volunteer medical students, who collected the data in challenging circumstances, and the generosity of the participants and their families for their individual contributions in these difficult times. We also acknowledge the support of Jeremy J Farrar, Nahoko Shindo, Devika Dixit, Nipunie Rajapakse, Piero Olliaro, Lyndsey Castle, Martha Buckley, Debbie Malden, Katherine Newell, Kwame O’Neill, Emmanuelle Denis, Claire Petersen, Scott Mullaney, Sue MacFarlane, Chris Jones, Nicole Maziere, Katie Bullock, Emily Cass, William Reynolds, Milton Ashworth, Ben Catterall, Louise Cooper, Terry Foster, Paul Matthew Ridley, Anthony Evans, Catherine Hartley, Chris Dunn, Debby Sales, Diane Latawiec, Erwan Trochu, Eve Wilcock, Innocent Gerald Asiimwe, Isabel Garcia-Dorival, J. Eunice Zhang, Jack Pilgrim, Jane A Armstrong, Jordan J. Clark, Jordan Thomas, Katharine King, Katie Alexandra Ahmed, Krishanthi S Subramaniam, Lauren Lett, Laurence McEvoy, Libby van Tonder, Lucia Alicia Livoti, Nahida S Miah, Rebecca K. Shears, Rebecca Louise Jensen, Rebekah Penrice-Randal, Robyn Kiy, Samantha Leanne Barlow, Shadia Khandaker, Soeren Metelmann, Tessa Prince, Trevor R Jones, Benjamin Brennan, Agnieska Szemiel, Siddharth Bakshi, Daniella Lefteri, Maria Mancini, Julien Martinez, Angela Elliott, Joyce Mitchell, John McLauchlan, Aislynn Taggart, Oslem Dincarslan, Annette Lake, Claire Petersen, and Scott Mullaney.

## Author Contributions

AMG - designed the study concept, reviewed all aspects of the data analysis and interpretation, contributed to the writing of the manuscript, and performed the final review of the manuscript.

AJS - performed the data analysis and contributed to the data interpretation and the writing of the manuscript.

SHK - performed the initial literature search and contributed to the data analysis and interpretation and the writing of the manuscript.

MC - contributed to conceptualisation and review of the manuscript

LW - contributed to conceptualisation and review of the manuscript

SC - contributed to conceptualisation and review of the manuscript

GV - contributed to data analysis and interpretation and the writing of the manuscript

AD - contributed to the ISARIC CCP-UK study design and data collection and reviewed the manuscript

EMH - contributed to the ISARIC-CCP UK study design and data collection and reviewed the manuscript

LT-contributed to the ISARIC-CCP UK study design and data collection and reviewed the manuscript

PJMO - ISARIC CCP-UK Co-Lead investigator, sourced funding, contributed to the

ISARIC-CCP-UK study design and data collection and reviewed the manuscript

JKB - ISARIC CCP-UK Consortium lead investigator, sourced funding, contributed to the

ISARIC CCP-UK study design and data collection and reviewed the manuscript

CAS - advised on all aspects of the conceptualisation and data analysis and interpretation, and contributed to the writing and final review of the manuscript

MGS - ISARIC CCP-UK Protocol Chief Investigator and guarantor of the data, sourced permissions and funding, contributed to the

ISARIC CCP-UK study design and data collection and reviewed the manuscript.

## ISARIC 4C Investigators

Consortium Lead Investigator: J Kenneth Baillie, Chief Investigator Malcolm G Semple Co-Lead Investigator Peter JM Openshaw. ISARIC Clinical Coordinator Gail Carson. Co-Investigators: Beatrice Alex, Benjamin Bach, Wendy S Barclay, Debby Bogaert, Meera Chand, Graham S Cooke, Annemarie B Docherty, Jake Dunning, Ana da Silva Filipe, Tom Fletcher, Christopher A Green, Ewen M Harrison, Julian A Hiscox, Antonia Ying Wai Ho, Peter W Horby, Samreen Ijaz, Saye Khoo, Paul Klenerman, Andrew Law, Wei Shen Lim, Alexander, J Mentzer, Laura Merson, Alison M Meynert, Mahdad Noursadeghi, Shona C Moore, Massimo Palmarini, William A Paxton, Georgios Pollakis, Nicholas Price, Andrew Rambaut, David L Robertson, Clark D Russell, Vanessa Sancho-Shimizu, Janet T Scott, Louise Sigfrid, Tom Solomon, Shiranee Sriskandan, David Stuart, Charlotte Summers, Richard S Tedder, Emma C Thomson, Ryan S Thwaites, Lance CW Turtle, Maria Zambon. Project Managers Hayley Hardwick, Chloe Donohue, Jane Ewins, Wilna Oosthuyzen, Fiona Griffiths. Data Analysts: Lisa Norman, Riinu Pius, Tom M Drake, Cameron J Fairfield, Stephen Knight, Kenneth A Mclean, Derek Murphy, Catherine A Shaw. Data and Information System Manager: Jo Dalton, Michelle Girvan, Egle Saviciute, Stephanie Roberts Janet Harrison, Laura Marsh, Marie Connor. Data integration and presentation: Gary Leeming, Andrew Law, Ross Hendry. Material Management: William Greenhalf, Victoria Shaw, Sarah McDonald. Outbreak Laboratory Volunteers: Katie A. Ahmed, Jane A Armstrong, Milton Ashworth, Innocent G Asiimwe, Siddharth Bakshi, Samantha L Barlow, Laura Booth, Benjamin Brennan, Katie Bullock, Benjamin WA Catterall, Jordan J Clark, Emily A Clarke, Sarah Cole, Louise Cooper, Helen Cox, Christopher Davis, Oslem Dincarslan, Chris Dunn, Philip Dyer, Angela Elliott, Anthony Evans, Lewis WS Fisher, Terry Foster, Isabel Garcia-Dorival, Willliam Greenhalf, Philip Gunning, Catherine Hartley, Antonia Ho, Rebecca L Jensen, Christopher B Jones, Trevor R Jones, Shadia Khandaker, Katharine King, Robyn T. Kiy, Chrysa Koukorava, Annette Lake, Suzannah Lant, Diane Latawiec, L Lavelle-Langham, Daniella Lefteri, Lauren Lett, Lucia A Livoti, Maria Mancini, Sarah McDonald, Laurence McEvoy, John McLauchlan, Soeren Metelmann, Nahida S Miah, Joanna Middleton, Joyce Mitchell, Shona C Moore, Ellen G Murphy, Rebekah Penrice-Randal, Jack Pilgrim, Tessa Prince, Will Reynolds, P. Matthew Ridley, Debby Sales, Victoria E Shaw, Rebecca K Shears, Benjamin Small, Krishanthi S Subramaniam, Agnieska Szemiel, Aislynn Taggart, Jolanta Tanianis, Jordan Thomas, Erwan Trochu, Libby van Tonder, Eve Wilcock, J. Eunice Zhang. Local Principal Investigators: Kayode Adeniji, Daniel Agranoff, Ken Agwuh, Dhiraj Ail, Ana Alegria, Brian Angus, Abdul Ashish, Dougal Atkinson, Shahedal Bari, Gavin Barlow, Stella Barnass, Nicholas Barrett, Christopher Bassford, David Baxter, Michael Beadsworth, Jolanta Bernatoniene, John Berridge, Nicola Best, Pieter Bothma, David Brealey, Robin Brittain-Long, Naomi Bulteel, Tom Burden, Andrew Burtenshaw, Vikki Caruth, David Chadwick, Duncan Chambler, Nigel Chee, Jenny Child, Srikanth Chukkambotla, Tom Clark, Paul Collini, Catherine Cosgrove, Jason Cupitt, Maria-Teresa Cutino-Moguel, Paul Dark, Chris Dawson, Samir Dervisevic, Phil Donnison, Sam Douthwaite, Ingrid DuRand, Ahilanadan Dushianthan, Tristan Dyer, Cariad Evans, Chi Eziefula, Chrisopher Fegan, Adam Finn, Duncan Fullerton, Sanjeev Garg, Sanjeev Garg, Atul Garg, Jo Godden, Arthur Goldsmith, Clive Graham, Elaine Hardy, Stuart Hartshorn, Daniel Harvey, Peter Havalda, Daniel B Hawcutt, Maria Hobrok, Luke Hodgson, Anita Holme, Anil Hormis, Michael Jacobs, Susan Jain, Paul Jennings, Agilan Kaliappan, Vidya Kasipandian, Stephen Kegg, Michael Kelsey, Jason Kendall, Caroline Kerrison, Ian Kerslake, Oliver Koch, Gouri Koduri, George Koshy, Shondipon Laha, Susan Larkin, Tamas Leiner, Patrick Lillie, James Limb, Vanessa Linnett, Jeff Little, Michael MacMahon, Emily MacNaughton, Ravish Mankregod, Huw Masson, Elijah Matovu, Katherine McCullough, Ruth McEwen, Manjula Meda, Gary Mills, Jane Minton, Mariyam Mirfenderesky, Kavya Mohandas, Quen Mok, James Moon, Elinoor Moore, Patrick Morgan, Craig Morris, Katherine Mortimore, Samuel Moses, Mbiye Mpenge, Rohinton Mulla, Michael Murphy, Megan Nagel, Thapas Nagarajan, Mark Nelson, Igor Otahal, Mark Pais, Selva Panchatsharam, Hassan Paraiso, Brij Patel, Justin Pepperell, Mark Peters, Mandeep Phull, Stefania Pintus, Jagtur Singh Pooni, Frank Post, David Price, Rachel Prout, Nikolas Rae, Henrik Reschreiter, Tim Reynolds, Neil Richardson, Mark Roberts, Devender Roberts, Alistair Rose, Guy Rousseau, Brendan Ryan, Taranprit Saluja, Aarti Shah, Prad Shanmuga, Anil Sharma, Anna Shawcross, Jeremy Sizer, Richard Smith, Catherine Snelson, Nick Spittle, Nikki Staines, Tom Stambach, Richard Stewart, Pradeep Subudhi, Tamas Szakmany, Kate Tatham, Jo Thomas, Chris Thompson, Robert Thompson, Ascanio Tridente, Darell Tupper - Carey, Mary Twagira, Andrew Ustianowski, Nick Vallotton, Lisa Vincent-Smith, Shico Visuvanathan, Alan Vuylsteke, Sam Waddy, Rachel Wake, Andrew Walden, Ingeborg Welters, Tony Whitehouse, Paul Whittaker, Ashley Whittington, Meme Wijesinghe, Martin Williams, Lawrence Wilson, Sarah Wilson, Stephen Winchester, Martin Wiselka, Adam Wolverson, Daniel G Wooton, Andrew Workman, Bryan Yates, Peter Young.

